# Ranked placement of phage predation as a determinant of dehydration severity among cholera patients in Bangladesh

**DOI:** 10.1101/2025.06.17.25329780

**Authors:** N. Madi, Md. Abu Sayeed, E. T. Cato, A. Creasy-Marrazzo, K. Islam, Md. I.UL. Khabir, Md. T. R. Bhuiyan, Y. Begum, E. Freeman, A. Vustepalli, L. Brinkley, M. Kamat, L. S. Bailey, K. B. Basso, F. Qadri, A. I. Khan, B. J. Shapiro, E. J. Nelson

## Abstract

Phage predation is inversely associated with severe cholera yet its importance as a determinant of dehydration severity is unknown relative to other factors. Here we used machine learning to assess and rank potential host, microbial, and environmental factors as determinants of severe dehydration among a cohort of cholera patients enrolled at hospital admission across Bangladesh. We found the phage to pathogen ratio ranked among the top classifying features, placing just behind patient age and admission location. We advocate that phage predation is a key factor to include in the characterization of cholera for scientific, clinical and epidemiological applications.

## MAIN TEXT

Cholera is an acute diarrheal disease caused by the Gram-negative bacterium *Vibrio cholerae* (*Vc*). Determinants of severe dehydration include diverse host, microbial, and environmental factors^1–3,4^. We previously found that predation by virulent bacteriophages on *Vc* inversely correlated with dehydration severity. Using a simple threshold of one phage (ICP1) to five *Vc*, we differentiated patients with and without dehydration (85% sensitivity; 53% specificity)^1^. While intriguing, the relative importance of phage compared to such factors as patient age, location, antibiotic exposure and pathogen abundance is unknown. Addressing this knowledge gap may be key to better characterizing cholera pathophysiology and informing clinical, diagnostic and public health strategies.

To address this gap, we applied multiple machine learning (ML) models in a secondary analysis of a prospective longitudinal study of cholera patients enrolled at hospital admission at seven hospitals across Bangladesh. Our objective was to identify and rank potential determinants of dehydration. We selected specific host demographic (age, gender, hospital location), microbial (*Vc* and phage abundance, phage:*Vc* ratios), and environmental (antibiotic exposure determined by mass spectrometry) factors as potential predictors of dehydration status. Our guiding hypothesis was that phage predation would rank high in importance in discriminating patients with and without dehydration.

## METHODS

### Ethics statement

The parent study that enabled the secondary analysis herein was led by AIK and FQ in Bangladesh who sought ERC/RRC approvals at the icddr,b and EJN who led the laboratory research at the University of Florida. BS conducted analyses on a deidentified dataset. Associated protocol numbers/registrations were IEDCR IRB/2017/10; icddr,b ERC/RRC PR-17036; University of Florida IRB 201601762; clinicaltrials.gov NCT03154229.

### Study design

This study represents a secondary analysis of a longitudinal prospective study of cholera patients with diarrheal disease who were admitted to one of seven hospitals across Bangladesh in a one-year period ^5^. We analyzed records of 623 cholera patients out of a cohort of 2462 patients.

### Dataset description

A cholera patient was defined microbiologically by culture positivity for *Vc*, molecular detection of *Vc,* or the *Vc-*specific bacteriophage (ICP1)^1, 5^. Among the 623 patients, 82 (13.2%) had ‘Mild’, 254 (40.8%) had ‘Moderate’ and 287 (46.1%) had ‘Severe’ dehydration by WHO measures. Mass spectrometry (LC-MS/MS) was performed on a subset of cholera patients (n=266) described previously ^1^ and analyzed herein. This dataset is referred to as the ‘small’ dataset, in contrast to the ‘large’ dataset of 623 patients. Of those samples analyzed by LC-MS/MS in the small dataset, 16 (6%) had ‘Mild’, 101 (38%) ‘Moderate’ and 149 (56%) ‘Severe’ dehydration. Quantitative polymerase chain reaction (qPCR) was performed on all the samples to determine absolute abundances of *Vc* and phages (ICP1/2/3)^1^.

### Machine learning analyses

Our first approach was to analyze a subset of variables as predictors of patients with ‘mild’, ‘moderate’, or ‘severe’ dehydration. Second, we repeated these analyses using a binary outcome of patients with dehydration (severe or moderate) and without dehydration (mild). Using the WHO conventions which have variable accuracy, patients with dehydration, or ‘non-mild’ dehydration, have at least 3-5% body weight loss^6^.

Analyses were conducted using seven machine learning (ML) algorithms from the scikit-learn Python library (version 1.4.1), Python 3.9.18. These included three implementations of gradient boosting decision tree models: Extreme Gradient Boosting (XGBoost), Light Gradient Boosting Machine (LightGBM) and Categorical Boosting (Catboost) ^7^. The other algorithms included the ensemble decision tree method, Random Forests (RF), the distance-based K-Nearest Neighbors (KNN), and two linear models: Linear Support Vector Machine (SVM) and Logistic Regression (LR). We applied one-hot encoding to categorical variables before training using the OneHotEncoder function from the feature_engine python package. While most classifiers require numeric input features, the CatBoost algorithm handles categorical variables natively without needing to convert them to numeric values before training.

To address class imbalance in our data, where moderate and severe dehydrated patients are more prevalent, we used cost-sensitive classifiers by setting the class weight to ‘balanced’. For classifiers such as logistic regression and CatBoost that do not accept this parameter, we calculated class weights using sklearn’s compute_class_weight module. The seven algorithms were optimized through hyperparameter tunning to identify the most effective parameter distribution and maximize the area under the curve of receiver operating characteristic plots (AUC-ROC). This was performed using the sklearn’s RandomizedSearchCV module which uses a dictionary of parameter distributions and runs a number (5) of cross-validaton parameter settings. Each run samples 10 different random combinations of parameters from the distributions, resulting in a total of 50 model fits to determine the best performing parameters.

We used the following hyperparameters for each algorithm: (i) KNN: k, metric, p, (ii) SVM: C, gamma, kernel, (iii) RF: bootstrap, criterion, max_depth, max_features, min_samples_leaf, min_samples_split, n_estimators, (iv) LGBM: boosting_type, colsample_bytree, importance_type, learning_rate, max_depth, min_samples_leaf, min_samples_split, n_estimators, num_leaves, reg_alpha, reg_lambda, subsample, (v) XGBC: gamma, learning_rate, n_estimators, reg_alpha, reg_lambda, (vi), LR: solver, penalty, max_iter, C, and (vii) Catboost: - boosting_type, colsample_bylevel, depth, iterations, l2_leaf_reg, learning_rate, min_data_in_leaf. For simplicity, we considered six predictors previously associated with dehydration severity. These predictors were: *Vc* and phage (ICP1) abundance from qPCR, the ICP1:*Vc* ratio, self-reported gender (male, female), age (7 months-75 years), and the hospital identity (7 different hospitals across Bangladesh). Additionally, for the smaller subset of patients with LC-MS/MS data, we included the concentrations, in μg/ml if detected, of three antibiotics commonly used to treat cholera (Azithromycin; AZI, Ciprofloxacin; CIP, and doxycycline; DOX).

The models were trained on 80% of the data and tested on the remaining 20% holdout set, using the Python package scikit-learn 1.4.1, in Python 3.9.18. To ensure that the proportion of each class was preserved in both the training and the testing sets, we used stratified split (using the stratify parameter in the train_test_split function). To assess performances, the classifiers were trained on each training dataset produced by the stratified 100-fold cross-validation (CV) of the input dataset. CV can assess a model’s performance more robustly and reduce the risk of overfitting to a single train-test split especially when the data is imbalanced, as is the case here. Stratified CV ensures that each fold maintains the same proportion of classes as the original dataset. By randomly sampling from the majority and minority classes according to the original distribution, we ensure more robust validation, as the distributions of the partitions will be similar to the initial distribution. Stratified CV is recommended when data is imbalanced because it provides a more accurate performance evaluation across all classes^8, 9^.

We compared the performance of the different algorithms by evaluating the average ROC-AUC from 100-fold cross-validation. AUC measures the model’s ability to distinguish between classes and is particularly useful for evaluating prediction performance on imbalanced data as it is invariant to class imbalance^10, 11^. We also reported other standard evaluation metrics like F1 score, precision, recall, and accuracy. Because our multi-class models have a class-imbalance, we adapted the AUC for multi-class classification by using the one-versus-rest (ovr) strategy, along with weight-averaging in Python’s sklearn.metrics library. All the other metrics were averaged using weights. We reported the feature importance for the best-performing model based on weighted AUC-ROC.

In the final set of analysis, we identified and ultimately ranked features contributing most to the classification, we calculated their importance using the Shapley Additive Explanation (SHAP) method^12^. SHAP (Shap 0.44.0 package in Python) is an approach exploring output from ML models using game theory. It provides a way to understand the contribution of each feature to the classifier’s prediction.

## RESULTS

We applied seven ML algorithms as multi-class (‘mild’, ‘moderate’, and ‘severe’) or binary classifiers (‘mild’ vs. ‘non-mild’). We trained each classifier on nine different features against a ‘small’ and ‘large’ dataset; the small dataset was restricted to patient samples matched with LC-MS/MS data. Catboost and Random Forests (RF) performed best across all metrics in both multi-class classifications and binary (Table S1). Both classifiers showed similar performance and achieved similarly high AUCs, a metric known to be invariant to class imbalance^10, 11^. RF achieved the highest AUC in the multi-class (0.801; 95% CI: 0.793-0.809) and the binary (0.948; 95% CI:0.939-0.958) models when trained on the small dataset. In contrast, Catboost outperformed RF in both the multi-class (0.825; 95% CI: 0.82-0.83) and the binary models (0.955; 95% CI: 0.951-0.958) when trained on the large dataset lacking antibiotic data (Table S1). Binary models showed improvements in all performance metrics for the prediction of patients with and without dehydration (Table S1). These two models generally agreed in their ranking of feature importance.

Across all models and datasets, sampling location and age were identified as the top two most important predictors of dehydration severity (Figures 1 and 2; Figures S1-S4). Among microbial features, the multi-class RF model applied to the large dataset revealed three key findings: (i) absolute *Vc* abundance was the strongest positive determinant for severe dehydration; (ii) ICP1 was the strongest negative determinant of moderate dehydration; (iii) The ratio of phage (ICP1) to *Vc* was the strongest positive feature discriminating patients with and without dehydration (Figure 2; Figure S2). Catboost yielded similar results except that low *Vc* abundance was more important than ICP1 or the ICP1:*Vc* ratio in predicting non-dehydrated patients (Figures S3, S4).

**Figure 1.**
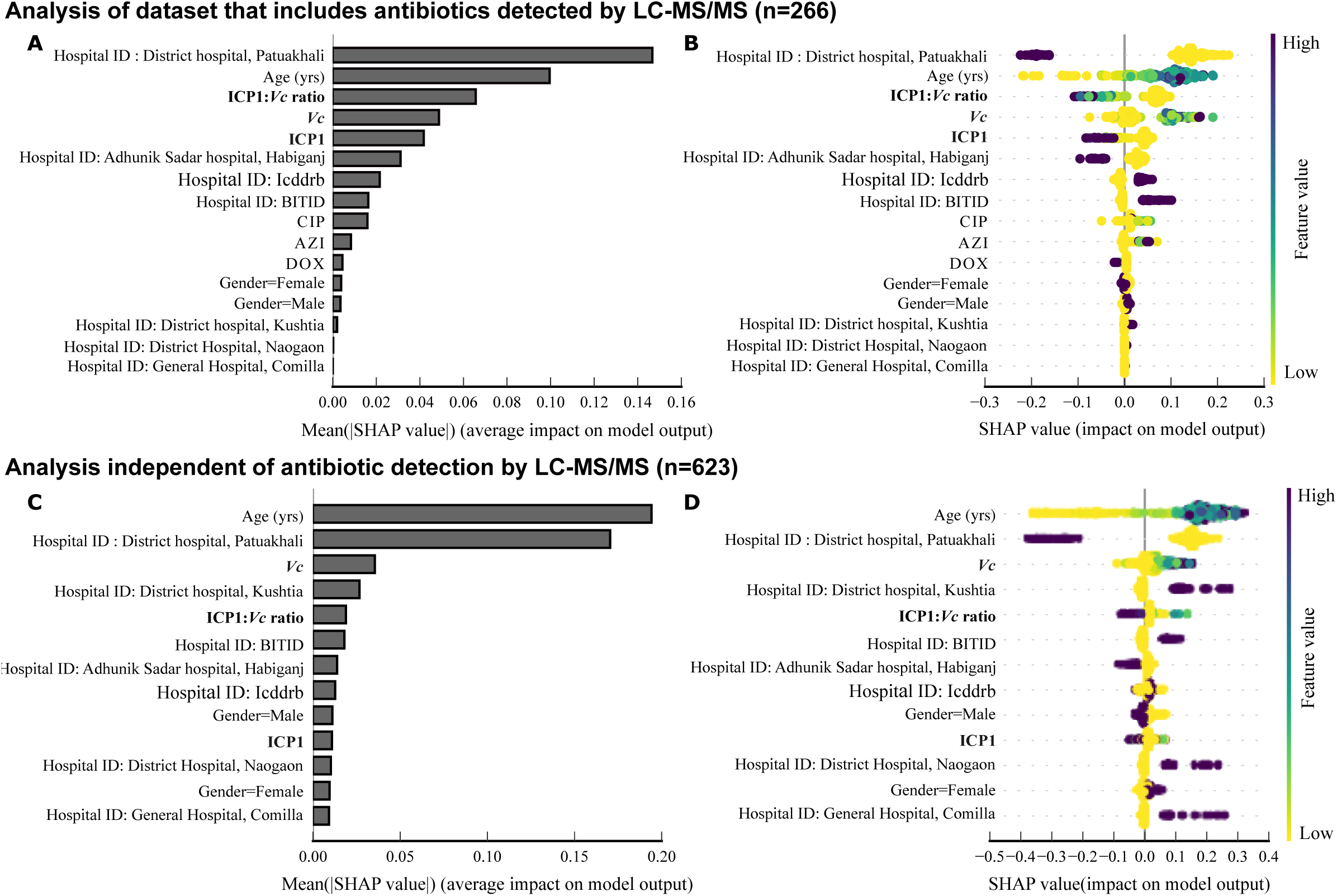
Binary feature importance in predicting ‘non-mild’ dehydration from the RF model trained on datasets that include (A, B) or are independent (C, D) of antibiotic quantification by LC-MS/MS. A, C: The importance ranking of features according to the average absolute impact (absolute mean SHAP values). B, D: Directional relationships between each patient feature and dehydration severity. Positive SHAP values indicate features that increase the prediction for non-mild status, and negative SHAP values indicate features that decrease the prediction of non-mild status. Larger SHAP values indicate a greater influence on the model predictions. Each point represents a single patient. Y-axis: features ordered by their importance in predicting non-mild dehydration. X-axis: SHAP values indicating magnitude and direction of influence. Among the smaller dataset that includes antibiotic detection by LC-MS/MS in panel B, RF was the best-performing binary model trained on the small dataset with an AUC-ROC of 0.948 (95% CI: 0.939-0.958). The model was trained on 266 patients, 16 (6%) mild and 250 (94%) non-mild, and nine features (*Vc*, ICP1, ICP1:*Vc*, AZI, CIP, DOX, hospital ID, patient’s age and self-reported gender). Among the larger dataset that excludes antibiotic detection by LC-MS/MS in panel D, RF was the second-best performing binary model trained on the large dataset with an AUC-ROC of 0.95 (95% CI: 0.946-0.954). The model was trained on 623 patients (82 mild, 541 non-mild) and six features (*Vc*, ICP1, ICP1:*Vc*, patient’s age and gender and hospital ID). Microbial features (*Vc,* ICP1, and their ratio) are shown in bold. Catboost model results are shown in the supplementary materials.

**Figure 2.**
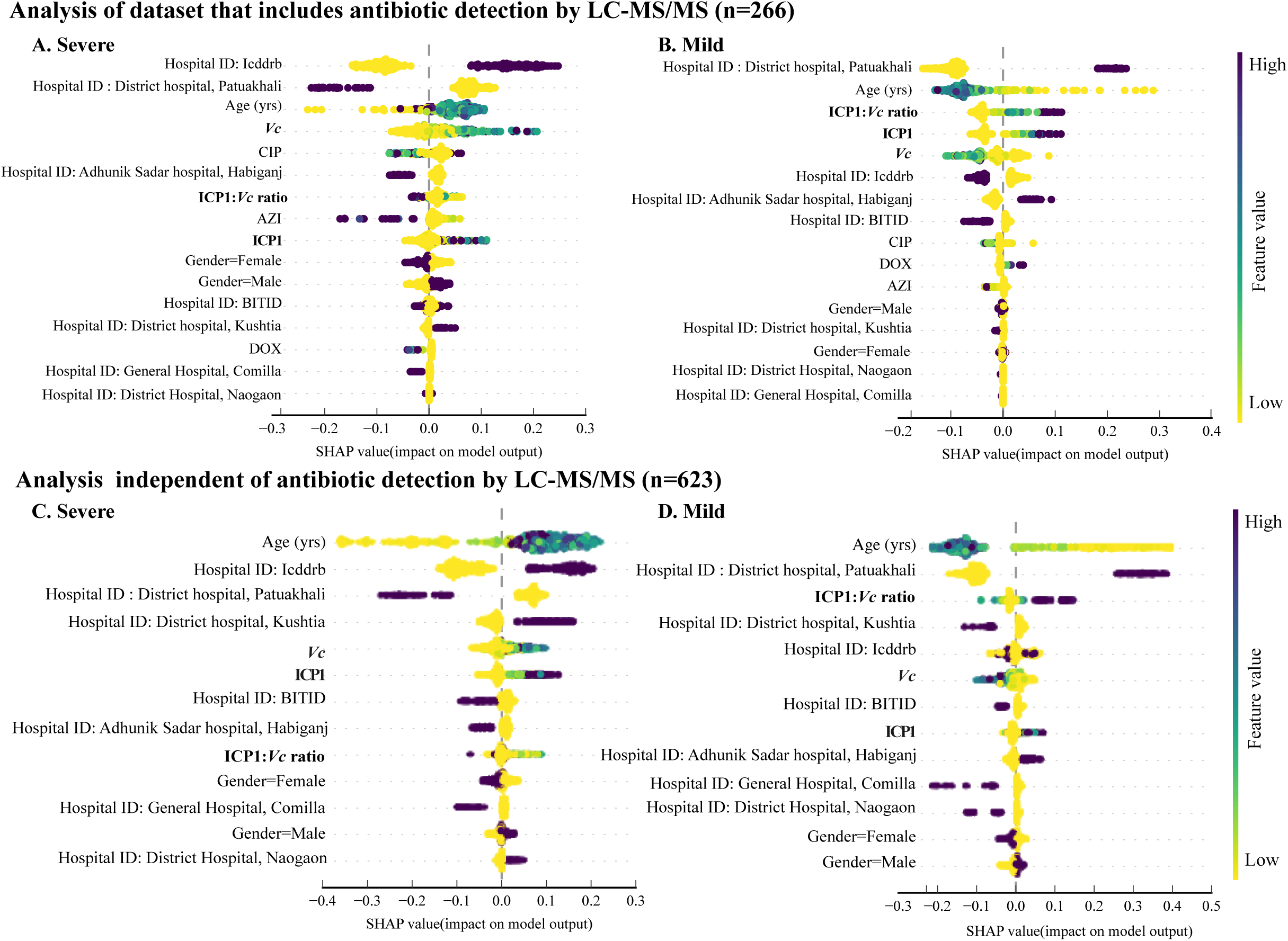
Multi-class feature importance of the RF model trained on datasets that include or are independent of antibiotic quantification by LC-MS/MS. Directional relationships between each patient feature and (A, C) severe or (B, D) mild dehydration. Positive SHAP values indicate features that increase the prediction, and negative SHAP values indicate features that decrease the prediction. Larger SHAP values indicate a greater influence on the model predictions. Each point represents a single patient. Y-axis: features ordered by their importance in the prediction. X-axis: SHAP values indicating magnitude and direction of influence. A) RF was the best-performing multi-class model trained on the small dataset with an AUC-ROC of 0.801 (95% CI: 0.793-0.809). This model was trained on 266 patients (16 mild, 101 moderate and 149 Severe) and nine features (*Vc*, ICP1, ICP1:*Vc*, AZI, CIP, DOX, hospital ID, patient’s age and gender). B) RF was the second-best performing multi-class model trained on the dataset excluding antibiotics with an AUC-ROC of 0.821 (95% CI: 0.816-0.826). The model was trained on 623 patients (82 mild, 254 moderate and 287 Severe) and six features (*Vc*, ICP1, ICP1:*Vc*, patient’s age and gender and hospital ID). Microbial features (*Vc,* ICP1, and their ratio) are shown in bold. Catboost model results are shown in the supplementary materials.

In the binary model applied to the smaller dataset, RF identified the ICP1:*Vc* ratio as the third most important feature in predicting patients with or without dehydration (Figure 1A-B). In contrast, the ratio had less impact than *Vc* alone when the large dataset was used (Figure 1C-D). While some of this difference may be attributable to small sample size, it is also possible that the inclusion of antibiotic exposure data affects the ranking of other predictors. With Catboost, *Vc* and the ratio had the same direction of disease association, but their effects were negligible for the large and small datasets (Figures S3, S4). Of the three antibiotics measured, CIP and AZI had intermediate effects in the RF multi-class model (Figure 2A-B; Figure S1). Antibiotics had minimal effects on dehydration status in the binary model (Figure 1A-B).

## DISCUSSION

In this study, we aimed to evaluate and rank phage predation as a determinant of dehydration severity among cholera patients in Bangladesh. To address this objective we leveraged ML-based approaches to evaluate the relative importance of phage predation against known demographic determinants^3, 4, 13, 14^ ^2,15–18^. This objective is important because phage predation was a known determinant, but we lacked insight on its relative importance^1^. Taken together, we found the phage to pathogen ratio ranked among the top predictive features of dehydration, placing just behind patient age and admission location.

Our first approach involved a larger dataset that increased power to test hypotheses; however, some samples lacked LC-MS/MS data on antibiotics. The second approach consisted of a smaller dataset in which all samples had LC-MS/MS data. After hospital location and age, the multi-class models ranked phage predation (ICP1:*Vc* ratios) as the best predictor of mild dehydration, *Vc* as the best predictor of severe dehydration, and ICP1 as the best predictor of moderate dehydration, regardless of whether antibiotics were included in the models. In the binary models, the ICP1:*Vc* ratio was the best microbial predictor of mild dehydration when antibiotic data was included, whereas *Vc* was a slightly better predictor than the ratio in the larger cohort without antibiotic data. This could be due to the lack of antibiotic data masking the effect of ICP1 in the large dataset, or the effect of ICP1 being overestimated in the small dataset. Future studies will be improved by conducting LC-MS/MS on all samples given that patient self-reports of antibiotic usage are of limited value.

Applications of these findings may impact our fundamental understanding of the pathophysiology of cholera and related diarrheal diseases, diagnostic development and epidemiologic surveillance. While phage predation has been documented for over a century, considering phage predation in disease frameworks may help explain why seemingly identical patients have dramatically different clinical courses. Diagnostics that include phage detection might be improved by using phage as a proxy for pathogen detection when the phage is pathogen-specific and has degraded the primary pathogen. Lastly, improved diagnostics will in turn improve epidemiologic surveillance. Future validation of this research, including cost analyses, is needed to before implementation.

These findings need to be considered in the context of the study limitations. First, the sample size was relatively small and unbalanced. We conducted the analysis with the ‘large’ and ‘small’ datasets while acknowledging that both approaches had complementary strengths and weaknesses. Second, relatively few features were included in the analysis to avoid overfitting the models; these features were based on prior literature yet may have biased the analysis by unknowingly excluding potentially important features. Third, the samples analyzed were collected at hospital admission alone. Given that infection, phage predation, and transmission are dynamic processes, future sampling of patients over time would be informative. Despite these limitations, this study ranks phage predation for the first time within the fold of known factors associated with dehydration among cholera patients. The strengths and limitations of this study should inspire further research to test our core hypotheses for both cholera and potentially other infections that could be influenced by phage predation.

## Data Availability

Sequencing data were previously deposited in the NCBI SRA under BioProject PRJNA976726.
Code for figures and results in this study are available on Github at https://github.com/Naima16/cholera_severity_determinants

## Acknowledgements

We thank the patients for participating in the parent study as well as the teams who collected and processed the samples. We are grateful to the Institute of Epidemiology, Disease Control and Research (IEDCR), Ministry of Health and Family Welfare, Government of Bangladesh who collaborated on the original clinical studies. Drs. C. Schmid and A. Levine at Brown University provided essential feedback that improved the manuscript. N. Rushing, T. Linn, and K. Berquist provided essential administrative support at the University of Florida.

## Author contributions

Conceptualization: NM, BJS, EJN

Methodology: NM, MAS, ETC, ACM, MIUK, MTRB, YB, EF, AV, LB, MK, LSB, KBS, BJS, EJN

Investigation: NM, BJS, EJN

Visualization: NM, BJS

Funding acquisition: KBS, FQ, AIK, BJS, EJN

Project administration: MTRB, YB, KBS, FQ, AIK, BJS, EJN

Supervision: BJS, EJN

Validation: NM Formal analysis: NM

Resources: BJS, AIK, EJN

Data Curation: NM

Writing – original draft: NM, BJS, EJN

Writing – review & editing: NM, AIK, BJS, EJN

## Data and materials availability

Sequencing data were previously deposited in the NCBI SRA under BioProject PRJNA976726.

## Code availability

Code for figures and results in this study are available on Github (https://github.com/Naima16/cholera_severity_determinants).

## SUPPLEMENTARY MATERIALS

### SUPPLEMENTARY FIGURES

**Figure S1.**
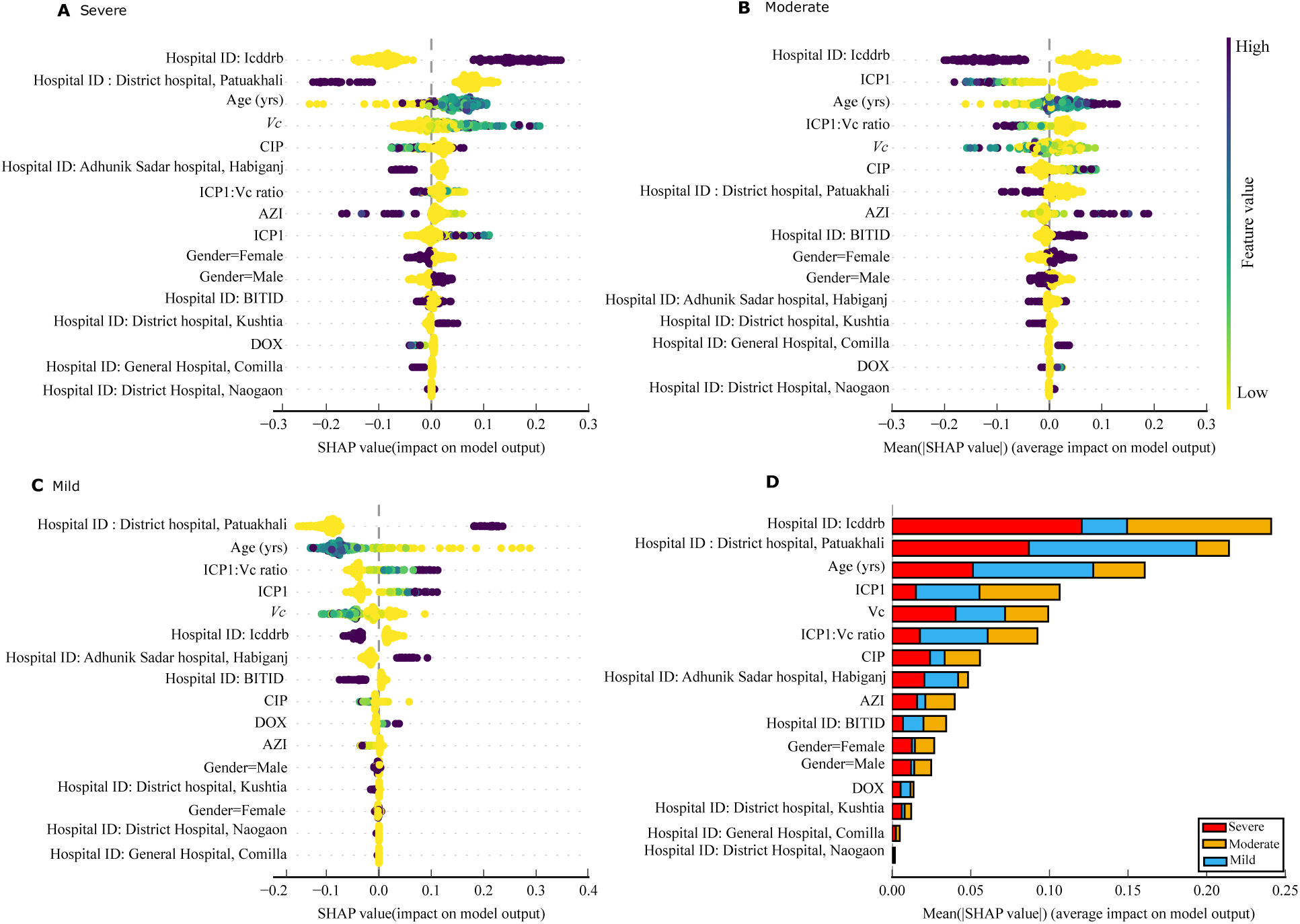
Multi-class feature importance of the RF model trained on the small dataset that includes antibiotic quantification. Directional relationships between each patient feature and A) severe, B) moderate and C) mild dehydration (A and C are also shown in Figure 2). D) *T*he importance ranking of the features according to the average absolute impact (absolute mean SHAP values) over all samples. Positive SHAP values indicate features that increase the prediction, and negative SHAP values indicate features that decrease the prediction. Larger SHAP values indicate a greater influence on the model’s prediction. Each point represents a single patient. Y-axis: features ordered by their importance in the prediction. X-axis: SHAP values indicating magnitude and direction of influence. RF is the best-performing multi-class model trained on the small dataset with an AUC-ROC of 0.801 (95% CI: 0.793-0.809). This model was trained on 266 patients (16 mild, 101 moderate and 149 Severe) and nine features (*Vc*, ICP1, ICP1:*Vc*, AZI, CIP, DOX, hospital ID, patient’s age and gender). Microbial features (*Vc,* ICP1, and their ratio) are shown in bold.

**Figure S2.**
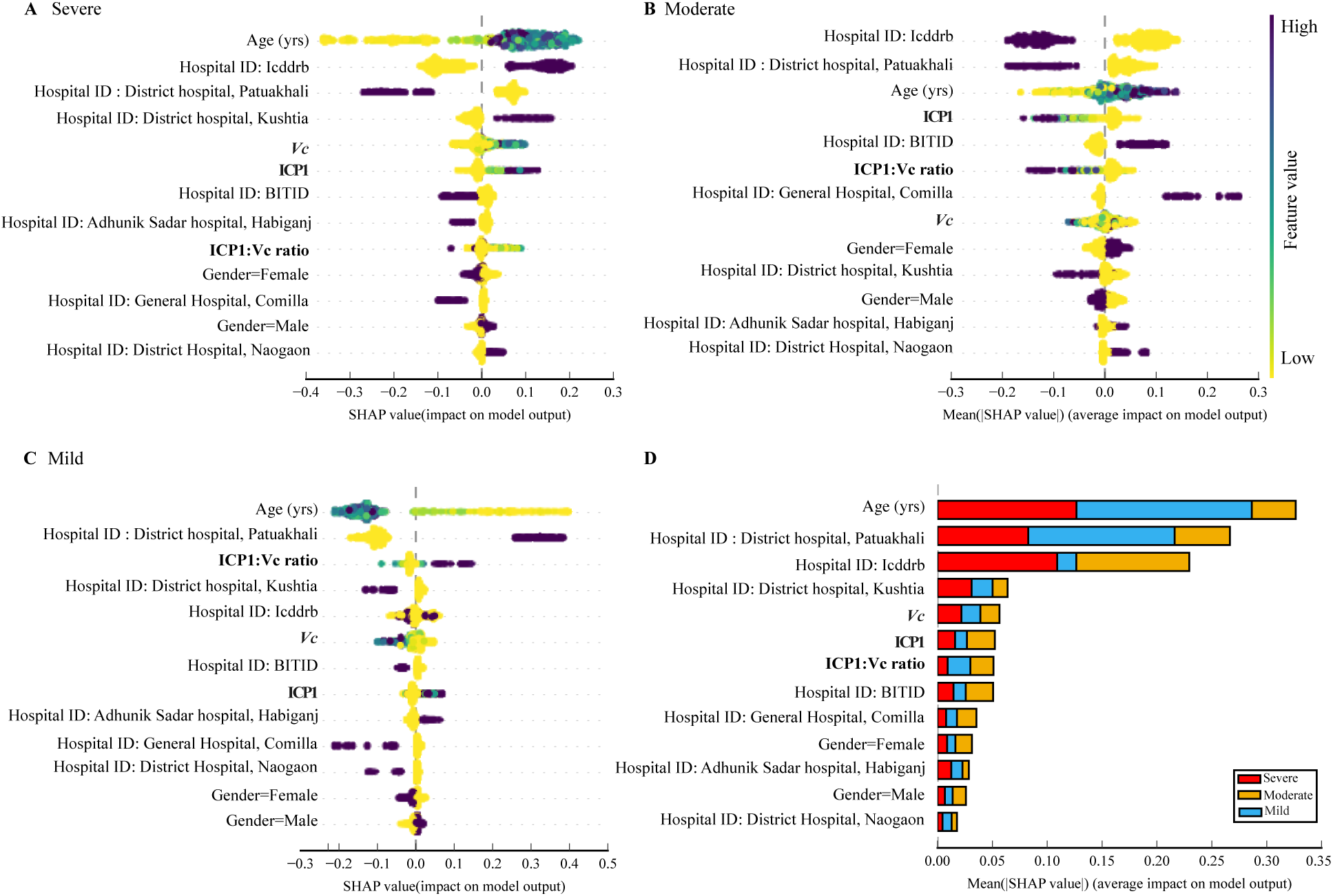
Multi-class feature importance from the RF model trained on the large dataset (antibiotic independent). Directional relationships between each patient feature and A) severe, B) moderate and C) mild dehydration (A and C are also shown in Figure 2). D) The importance ranking of the features according to the average absolute impact (absolute mean SHAP values) over all samples. Positive SHAP values indicate features that increase the prediction for non-mild status, and negative SHAP values indicate features that decrease the prediction of non-mild. Larger SHAP values indicate a greater influence on the model’s prediction. Each point represents a single patient. Y-axis: features ordered by their importance in the prediction. X-axis: SHAP values indicating magnitude and direction of influence. RF is the second-best performing multi-class model trained on the dataset excluding antibiotics with an AUC-ROC of 0.821 (95% CI: 0.816-0.826). The model was trained on 623 patients (82 mild, 254 moderate and 287 Severe) and six features (*Vc*, ICP1, ICP1:*Vc*, patient’s age and gender and hospital ID). Microbial features (*Vc,* ICP1, and their ratio) are shown in bold. Catboost model results are shown in Figure S1.

**Figure S3.**
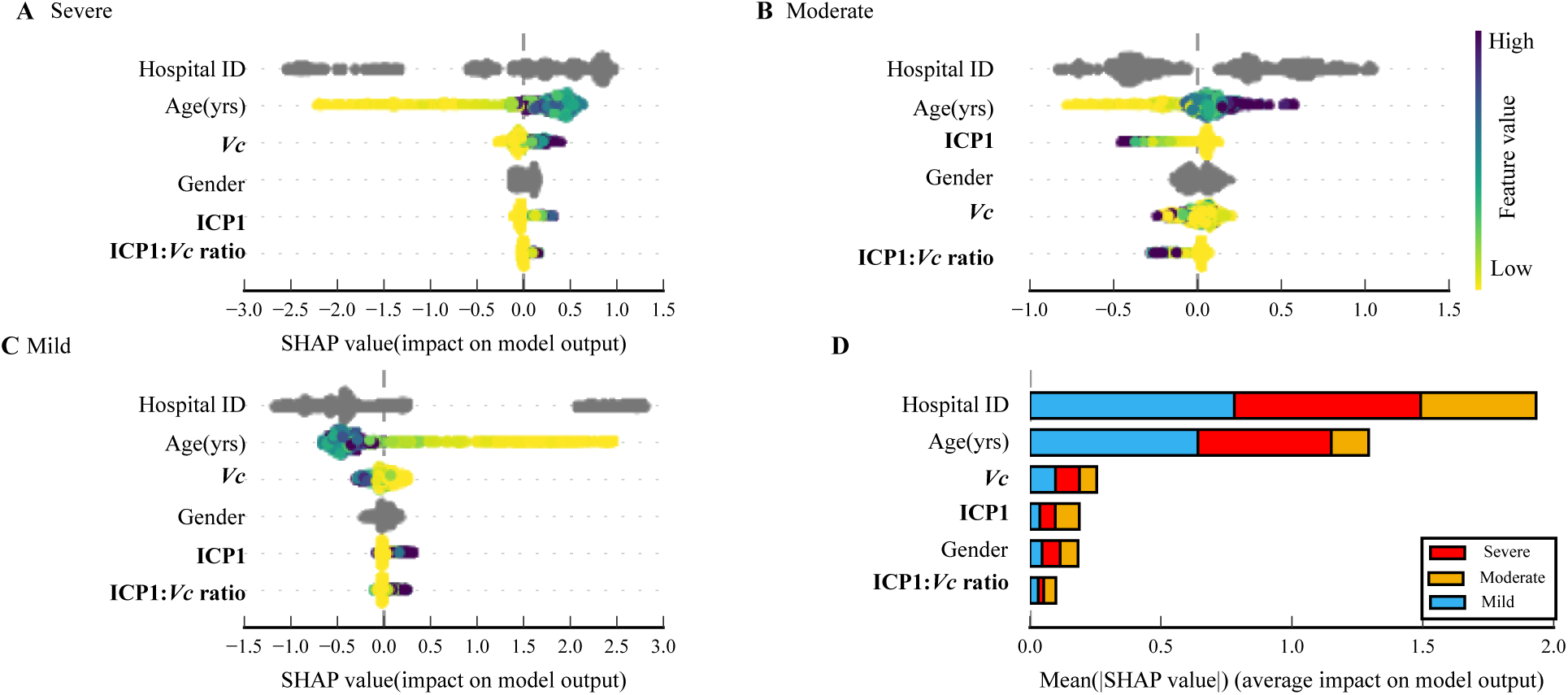
Multi-class feature importance from the catboost model trained on the large dataset (antibiotics independent). Directional relationships between each patient feature and A) severe, B) moderate and C) mild dehydration. D) The importance ranking of the features according to the average absolute impact (absolute mean SHAP values) over all samples. Positive SHAP values indicate features that increase the prediction for non-mild status, and negative SHAP values indicate features that decrease the prediction of non-mild. Larger SHAP values indicate a greater influence on the model’s prediction. Each point represents a patient from the data. Y-axis: features ordered by their importance in predicting A) severe, B) moderate and C) mild dehydration. X-axis: SHAP values indicating magnitude and direction of influence. Categorical variables are in grey because they are not numeric. Catboost is the best-performing multi-class model trained on the dataset, excluding antibiotics, with an AUC-ROC of 0.825 (95% CI: 0.82-0.83). 623 patients (82 mild, 254 moderate and 287 Severe) and six features (*Vc*, ICP1, ICP1:*Vc*, patient’s age and gender and hospital ID). Microbial features (*Vc,* ICP1, and their ratio) are shown in bold.

**Figure S4.**
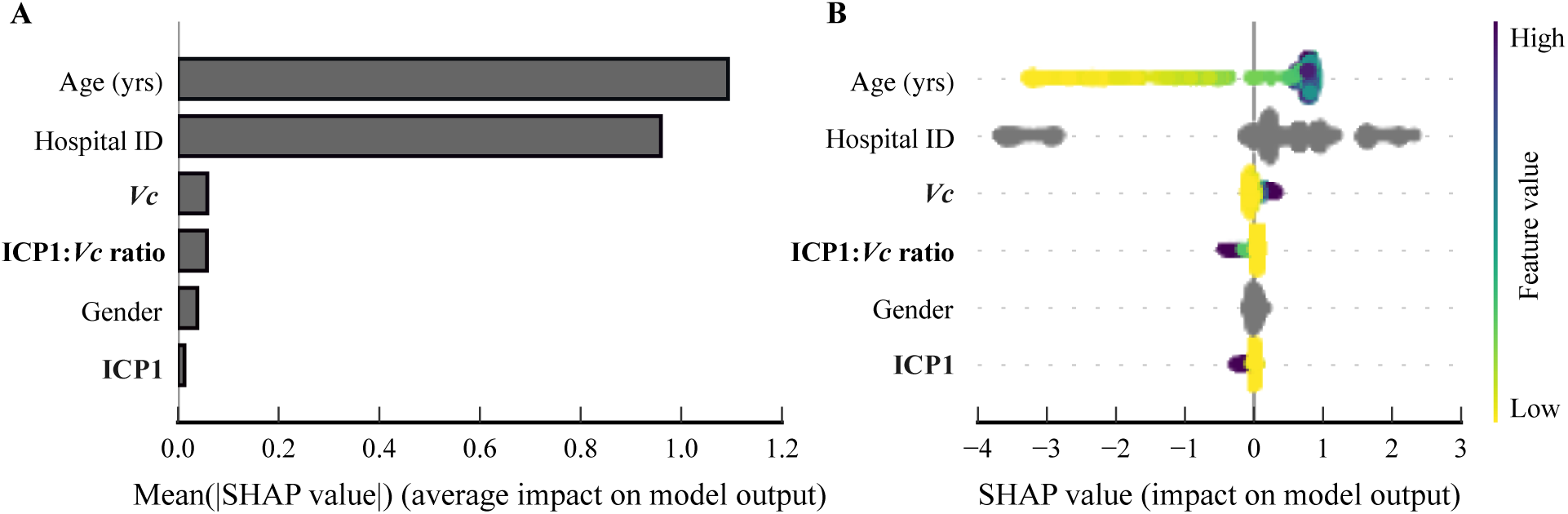
Binary feature importance in predicting ‘non-mild’ from the catboost model trained on the large dataset (antibiotics independent). A) The importance ranking of the features according to the average absolute impact (absolute mean SHAP values) over all samples. B) Directional relationships between each patient feature and dehydration severity. Positive SHAP values indicate features that increase the prediction for non-mild status, and negative SHAP values indicate features that decrease the prediction of non-mild. Larger SHAP values indicate a greater influence on the model’s prediction. Each point represents a patient from the data. Y-axis: features ordered by their importance in predicting non-mild dehydration. X-axis: SHAP values indicating magnitude and direction of influence. Categorical variables are in grey because they are not numeric. Catboost is the best-performing binary model trained on the dataset excluding antibiotics with an AUC-ROC of 0.955 (95% CI: 0.951-0.958). 623 patients, 82 (13.16%) mild and 541 (86.84%) non-mild and six features (*Vc*, ICP1, ICP1:*Vc*, patient’s age and gender and hospital ID). Microbial features (*Vc,* ICP1, and their ratio) are shown in bold.

### SUPPLEMENTARY TABLES

**Table S1.** Model performance across 100-fold cross-validation^a^.

|  |  | Small dataset (antibiotics measured) |  |  |  |  | Large dataset (antibiotics not measured) |  |  |  |  |
| --- | --- | --- | --- | --- | --- | --- | --- | --- | --- | --- | --- |
|  |  | ROC-AUC | F1 | recall | precision | acc | ROC-AUC | F1 | recall | precision | acc |
| Multi-class classifiers | RF | **0.801** | **0.697** | **0.697** | **0.709** | **0.697** | 0.821 | 0.681 | 0.682 | 0.694 | 0.682 |
|  |  | 0.793 | 0.688 | 0.687 | 0.699 | 0.687 | 0.816 | 0.674 | 0.675 | 0.687 | 0.675 |
|  |  | 0.809 | 0.707 | 0.706 | 0.718 | 0.706 | 0.826 | 0.688 | 0.689 | 0.7 | 0.689 |
|  | Catboost | 0.795 | 0.691 | 0.688 | 0.704 | 0.688 | **0.825** | **0.685** | **0.687** | 0.697 | **0.687** |
|  |  | 0.787 | 0.682 | 0.679 | 0.695 | 0.679 | 0.82 | 0.677 | 0.679 | 0.689 | 0.679 |
|  |  | 0.803 | 0.7 | 0.698 | 0.713 | 0.698 | 0.83 | 0.692 | 0.694 | 0.704 | 0.694 |
|  | LGBM | 0.759 | 0.648 | 0.649 | 0.658 | 0.649 | 0.785 | 0.639 | 0.64 | 0.643 | 0.64 |
|  |  | 0.75 | 0.638 | 0.639 | 0.647 | 0.639 | 0.779 | 0.631 | 0.632 | 0.635 | 0.632 |
|  |  | 0.768 | 0.658 | 0.659 | 0.668 | 0.659 | 0.791 | 0.647 | 0.648 | 0.65 | 0.648 |
|  | XGBoost | 0.767 | 0.596 | 0.639 | 0.617 | 0.639 | 0.819 | 0.684 | 0.686 | **0.707** | 0.686 |
|  |  | 0.758 | 0.581 | 0.631 | 0.599 | 0.631 | 0.814 | 0.677 | 0.678 | 0.699 | 0.678 |
|  |  | 0.776 | 0.611 | 0.647 | 0.635 | 0.647 | 0.825 | 0.692 | 0.694 | 0.715 | 0.694 |
|  | logistic | 0.784 | 0.691 | 0.69 | 0.703 | 0.69 | 0.813 | 0.678 | 0.679 | 0.69 | 0.679 |
|  |  | 0.776 | 0.682 | 0.681 | 0.694 | 0.681 | 0.807 | 0.671 | 0.672 | 0.683 | 0.672 |
|  |  | 0.793 | 0.701 | 0.7 | 0.712 | 0.7 | 0.819 | 0.686 | 0.687 | 0.698 | 0.687 |
|  | svm | 0.778 | 0.65 | 0.642 | 0.679 | 0.642 | 0.801 | 0.678 | 0.68 | 0.695 | 0.68 |
|  |  | 0.769 | 0.64 | 0.632 | 0.669 | 0.632 | 0.795 | 0.67 | 0.672 | 0.688 | 0.672 |
|  |  | 0.787 | 0.66 | 0.652 | 0.688 | 0.652 | 0.807 | 0.685 | 0.687 | 0.703 | 0.687 |
|  | knn | 0.586 | 0.596 | 0.639 | 0.617 | 0.639 | 0.532 | 0.684 | 0.686 | **0.707** | 0.686 |
|  |  | 0.575 | 0.581 | 0.631 | 0.599 | 0.631 | 0.526 | 0.677 | 0.678 | 0.699 | 0.678 |
|  |  | 0.596 | 0.611 | 0.647 | 0.635 | 0.647 | 0.539 | 0.692 | 0.694 | 0.715 | 0.694 |
| Binary classifiers | RF | **0.948** | **0.95** | **0.954** | 0.951 | **0.954** | 0.95 | **0.913** | **0.909** | 0.922 | **0.909** |
|  |  | 0.939 | 0.946 | 0.95 | 0.946 | 0.95 | 0.946 | 0.91 | 0.906 | 0.919 | 0.906 |
|  |  | 0.958 | 0.954 | 0.958 | 0.956 | 0.958 | 0.954 | 0.916 | 0.912 | 0.925 | 0.912 |
|  | Catboost | 0.933 | 0.941 | 0.934 | **0.954** | 0.934 | **0.955** | 0.904 | 0.894 | **0.935** | 0.894 |
|  |  | 0.916 | 0.935 | 0.927 | 0.949 | 0.927 | 0.951 | 0.901 | 0.89 | 0.933 | 0.89 |
|  |  | 0.951 | 0.947 | 0.942 | 0.958 | 0.942 | 0.958 | 0.908 | 0.898 | 0.937 | 0.898 |
|  | LGBM | 0.92 | 0.951 | 0.953 | 0.953 | 0.953 | 0.922 | 0.895 | 0.896 | 0.896 | 0.896 |
|  |  | 0.903 | 0.946 | 0.949 | 0.948 | 0.949 | 0.917 | 0.891 | 0.892 | 0.892 | 0.892 |
|  |  | 0.937 | 0.955 | 0.957 | 0.958 | 0.957 | 0.927 | 0.899 | 0.9 | 0.9 | 0.9 |
|  | XGBoost | 0.935 | 0.932 | 0.95 | 0.927 | 0.95 | 0.942 | 0.905 | 0.919 | 0.922 | 0.919 |
|  |  | 0.92 | 0.928 | 0.948 | 0.919 | 0.948 | 0.938 | 0.902 | 0.917 | 0.919 | 0.917 |
|  |  | 0.95 | 0.936 | 0.953 | 0.934 | 0.953 | 0.946 | 0.909 | 0.922 | 0.925 | 0.922 |
|  | logistic | 0.82 | 0.903 | 0.88 | 0.943 | 0.88 | 0.889 | 0.892 | 0.88 | 0.923 | 0.88 |
|  |  | 0.803 | 0.897 | 0.872 | 0.94 | 0.872 | 0.882 | 0.888 | 0.875 | 0.92 | 0.875 |
|  |  | 0.838 | 0.909 | 0.888 | 0.946 | 0.888 | 0.896 | 0.896 | 0.884 | 0.926 | 0.884 |
|  | svm | 0.713 | 0.928 | 0.926 | 0.933 | 0.926 | 0.887 | 0.882 | 0.867 | 0.921 | 0.867 |
|  |  | 0.69 | 0.923 | 0.921 | 0.928 | 0.921 | 0.88 | 0.878 | 0.863 | 0.918 | 0.863 |
|  |  | 0.737 | 0.932 | 0.93 | 0.937 | 0.93 | 0.894 | 0.886 | 0.872 | 0.923 | 0.872 |
|  | knn | 0.739 | 0.907 | 0.927 | 0.888 | 0.927 | 0.561 | 0.805 | 0.833 | 0.786 | 0.833 |
|  |  | 0.718 | 0.905 | 0.924 | 0.885 | 0.924 | 0.55 | 0.801 | 0.83 | 0.782 | 0.83 |
|  |  | 0.761 | 0.909 | 0.93 | 0.891 | 0.93 | 0.572 | 0.808 | 0.836 | 0.791 | 0.836 |

<sup>a</sup> The performance of seven different models are reported. Performance metrics correspond to the averaged AUC, F1, recall, precision, and accuracy values through 100-fold cross-validation, followed by the corresponding 95% confidence interval (lower and upper bounds). All estimates in the multi-class models are weighted by support (the metrics is calculated for each class and then the average is weighted by the number of instances in each class). The highest value for each metric is highlighted in black bold text.

